# A longitudinal study of SARS-CoV-2 infected patients shows high correlation between neutralizing antibodies and COVID-19 severity

**DOI:** 10.1101/2020.08.27.20182493

**Authors:** Vincent Legros, Solène Denolly, Manon Vogrig, Bertrand Boson, Josselin Rigaill, Sylvie Pillet, Florence Grattard, Sylvie Gonzalo, Paul Verhoeven, Omran Allatif, Philippe Berthelot, Carole Pélissier, Guillaume Thierry, Elisabeth Botelho-Nevers, Stéphane Paul, Thierry Walzer, François-Loïc Cosset, Thomas Bourlet, Bruno Pozzetto

## Abstract

Understanding the immune responses elicited by SARS-CoV-2 infection is critical in terms of protection from re-infection and, thus, for public health policy and for vaccine development against the COVID-19. Here, using either live SARS-CoV-2 particles or retroviruses pseudotyped with the SARS-CoV-2 S viral surface protein (Spike), we studied the neutralizing antibody (nAb) response in serum specimens from a cohort of 140 SARS-CoV-2 qPCR-confirmed patients, including patient with mild symptoms but also more severe form including those that require intensive care. We show that nAb titers were strongly correlated with disease severity and with anti-Spike IgG levels. Indeed, patients from intensive care units exhibited high nAb titers, whereas patients with milder disease symptoms displayed heterogenous nAb titers and asymptomatic or exclusive outpatient care patients had no or poor nAb levels. We found that the nAb activity in SARS-CoV-2-infected patients displayed a relatively rapid decline after recovery, as compared to individuals infected with alternative coronaviruses. We show the absence of cross-neutralization between endemic coronaviruses and SARS-CoV-2, indicating that previous infection by human coronaviruses may not generate protective nAb against SARS-CoV-2 infection. Finally, we found that the D614G mutation in the Spike protein, which has recently been identified as the major variant now found in Europe, does not allow neutralization escape. Altogether, our results contribute to the understanding of the immune correlate of SARS-CoV-2 induced disease and claim for a rapid evaluation of the role of the humoral response in the pathogenesis of SARS-CoV-2.

## Introduction

Emergence of SARS-CoV-2 was first detected in late 2019 in Wuhan, China. As per John Hopkins University and Coronavirus Resource Center, the SARS-CoV-2-induced disease, named COVID-19, has now caused by mid-August 2020 more than 750,000 reported deaths worldwide for over 21 million infected individuals, figures that are likely to be underestimated. The hallmark of the disease is an acute respiratory distress syndrome (ARDS), but other non-specific symptoms such as sore throat, dry cough, fever, fatigue, muscle aches, runny nose, and diarrhea are also frequently present [1]. Neurologic disorders have also been reported with headache, nausea, vomiting, anosmia and aguesia, acute cerebrovascular diseases, Guillain-Barré syndrome, and impaired consciousness [2].

Understanding the immune responses elicited by SARS-CoV-2 infection is critical in terms of protection from re-infection and, thus, for public health policy and vaccine development. One of the key roles of acquired immune responses is played by neutralizing antibodies (nAb) that are generally associated with virus clearance and protection [3, 4]. Several reports indicate that most of individuals recovering from SARS-CoV-2 infection develop IgM, IgG and IgA responses targeting the nucleocapsid (N) or the spike (S) protein of the SARS-CoV-2 viral particles 7–14 days after infection [5–7]. Additionally, nAbs have been identified in patients, suggesting that SARS-CoV-2 infection may generate a robust immune response [8, 9, 7]. Considering the lack of perspectives on the immune correlates of protection against SARS-CoV-2, it is tempting to draw conjecture from the immune responses elicited by other human coronaviruses. Hence, nAb activity of sera from patients infected with endemic coronaviruses can rapidly wane other time, as reinfection is frequently described [10], whereas nAbs against SARS-CoV and MERS-CoV can be detected for up to 36 months [11, 12]. It is therefore urgent to evaluate the nAb response elicited by SARS-CoV-2 infection, the factors associated with its robustness, and its persistence. Here, the nAb activity of serum specimens from a cohort of 140 SARS-CoV-2 qPCR-confirmed patients was quantified. We show that nAb titers are strongly correlated with disease severity. Importantly, we were able to quantify the persistence of nAb activity in patients, which indicated a relatively rapid decline of nAbs after recovery. We also showed the absence of cross-protection conferred by previous infection by endemic coronaviruses. Finally, we found that the D614G mutation in the Spike protein, that has recently been identified as the major variant now found in Europe [13] did not induce nAb escape.

## Methods

### Ethics

This study was approved by the Ethical Committee of the University Hospital of Saint-Etienne (reference number IRBN512020/CHUSTE).

### Patients and origin of samples

A total of 140 patients followed at the University Hospital of Saint-Etienne were enrolled between March and May 2020. All patients were sampled by nasal swab and tested positive for SARS-CoV-2 RNA by RT-qPCR assay. The patients were classified in 3 groups according to their medical care: 44 were admitted in intensive care unit (ICU), 42 were hospitalized (HOS) without receiving care in ICU, and 54 were given exclusive outpatient care (EOC) including 8 asymptomatic cases (ASY).

In the ICU and HOS groups, 3–4 sera were sampled at three periods of follow-up after the onset of symptoms: 0 to 15, 16 to 30 and > 30 days. In the EOC group, 2 sera were sampled from 13 to 62 days after the onset of symptoms.

Time post-onset was defined as the time after onset of the first symptoms.

### Seroneutralization assay using wild type SARS-CoV-2

The viral strain (RoBo strain) used this assay was a clinical isolate cultured on Vero-E6 cells (kind gift from Pr. B. La Scola, IHU Méditerranée, Marseille, France) from a nasopharyngeal aspirate of a patient hospitalized at the University Hospital of Saint-Étienne for severe COVID-19 infection; it was diluted in DMEM containing 2% FCS so that to obtain 100 to 500 tissue culture infectious doses 50% (TCID_50_) per ml. Each serum specimen was diluted 1:10 and serial two-fold dilutions were mixed with an equal volume (100 µl each) of the virus in a plastic microplate. After gentle shaking and a contact of 30 minutes at room temperature, 150 µl of the mixt was transferred to 96-well microplates covered with VeroE6 cells. The plates were then placed at 37°C in a 5% CO_2_ incubator. The reading was evaluated microscopically 5 to 6 days later when the cytopathic effect of the virus control reached about 100 TCID_50_/150 µl. The serum was considered to having protected the cells if more than 50% of the cell layer was preserved. The neutralizing titer was expressed as the inverse of the higher serum dilution that protected the cells.

### SARS-CoV-2 pseudoparticles preparation and determination of neutralization

SARS-CoV-2 Spike-pseudotyped murine leukemia virus (MLV) retrovirus particles were produced as described for SARS-CoV-1 [14]. Briefly, HEK293T cells (ATCC CRL-1573) were transfected with constructs expressing MLV Gag-Pol, the green fluorescent protein (GFP) reporter, and the SARS-CoV-2 Spike (kind gift from D. Lavillette). The plasmid encoding the Spike protein harboring D614G mutation was generated by PCR mutagenesis. Control pseudoparticles pseudotyped with the unrelated RD114 viral surface glycoprotein (from a cat endogenous virus) were generated as previously described [15]. For neutralization assays, around 1×10^3^ pseudoparticles were incubated with 100-fold dilution of sera or control antibodies 1h at 37°C prior to infection of VeroE6R (ATCC CRL-1586) cells. At 72h post transduction, the percentage of GFP-positive cells was determined on by flow cytometry. As control the same procedure was done using RD114 pseudoparticles.

Anti-Spike RBD SARS-CoV2 (Sino Biological) and anti-gp70 RD114 (79S914, ViroMed Biosafety Labs) were used as positive and negative control neutralizing antibodies, respectively.

### Commercial kits for measuring IgG antibodies against SARS-CoV-2

Two commercially-available kits were used for measuring the anti-SARS-CoV-2 IgG antibodies. The LIAISON® SARS-CoV-2 S1/S2 kit (Diasorin) measures antibodies against S1-S2 proteins whereas the ARCHITECT SARS-CoV-2 IgG kit (Abbott Laboratories) measures antibodies to the viral nucleoprotein.

### Statistical analysis

Analysis were performed using the GraphPad Prism 6 software. Significance values were calculated by applying the Kruskal-Wallis test and Dunn’s multiple comparison test or t-test depending on the population and Spearman’s coefficient and p-value were calculated to evaluate the correlation between variables. Second order polynomial regression were plotted by line and ribbons depicting the 95% confidence intervals. Parameters of the polynomial equations are indicated on the graphs. P values under 0.05 were considered statistically significant and the following denotations were used: ****, P<0.0001; ***, P<0.001; **, P<0.01; *, P<0.05; ns (not significant), P>0.05.

## Results

### COVID-19 patients and clinical information

One hundred and forty (140) laboratory-confirmed SARS-CoV-2 consenting patients from the University Hospital of Saint-Etienne (France) were enrolled in the study. Among them, 44 patients were admitted in intensive care unit (ICU), 42 patients were hospitalized (HOS) without receiving care in ICU, and 54 patients were given exclusive outpatient care (EOC) including 8 patients who were asymptomatic (ASY). Finally, 9 serum specimens from subjects infected by seasonal coronaviruses were also available for the study. 269 blood samples were collected from the patients at various time points post-onset of COVID-19 symptoms, based on the availability of discarded blood samples collected for routine clinical management. Importantly, several patients were sampled at least three times: 19 ICU, 14 HOS and 4 EOC, to up to 63 days after onset.

Overall, this cohort recapitulated the variability in the severity of COVID-19, and common comorbidities already identified [16-18] were also present in this cohort. A full summary of the characteristic of the patients is presented in Table 1. The main clinical symptoms in ICU and HOS groups were fever (90.1%), dyspnea (56.3%), cough (53.5%), asthenia (29.6%), diarrhea (23.9%), myalgia (8.5%), sputum production (7.0%) and anosmia/agueusia (4.2%). There were no differences of clinical symptoms between these two groups, which differed essentially with their severity. In the EOC group, moderate symptoms were mainly fever, cough and asthenia.

**Table 1.** Characteristics of the three groups of COVID-19 patients.

| **Characteristics** | **Patient's group** |  |  |
| --- | --- | --- | --- |
|  | **Intensive care**<br>**ICU (*n*=44)** | **Hospitalized**<br>**HOS (*n*=42)** | **Ambulatory**<br>**EOC + ASY**<br>**(*n*=54)** |
| **M/F sex ratio** | 0.6 | 1.6 | 3.9 |
| **Age (years), median (IQR)** | 70 (64-75.8) | 82 (65-85.2) | 37 (32-50.5) |
| **Comorbidities (%)** |  |  |  |
| Age >70 | 52.3 | 69.0 | 7.4 |
| Obesity | 25.0 | 9.7 | 0 |
| Hypertension | 62.5 | 61.3 | 0 |
| Diabetes | 30.0 | 22.6 | 0 |
| Cardiovascular disease | 12.5 | 41.9 | 0 |
| Kidney failure | 12.5 | 6.4 | 0 |
| Chronic respiratory diseases | 17.5 | 16.1 | 0 |
| Malignancy | 5 | 19.4 | 0 |
| **Severity criteria (%)** |  |  |  |
| Respiratory rate >30/min | 70 | 6.5 | 0 |
| Blood oxygen saturation | 97.5 | 22.6 | 0 |
| <92% | 92.5 | 38.7 | 0 |
| Severe injuries by CT scan | 100 | 45.1 | 0 |
| Oxygen and/or mechanical<br>ventilation needs |  |  |  |
| **Deceased (%)** | 16.3 | 15.4 | 0 |

### Pseudoparticles neutralization is correlated with wild-type SARS-CoV-2 neutralization

Neutralization of live virus in *in vitro* assays is considered as the gold-standard method for the assessment of nAbs. However, for SARS-CoV-2, it requires BSL-3 facility and it is time-consuming. Hence, we developed a SARS-CoV-2 pseudoparticle assay, named SARS-CoV-2pp, to quickly, safely and reliably assess nAb activity. This assay was compared to a classic neutralization assay performed with wild-type SARS-CoV-2 in BSL-3 facility. To identify unspecific neutralizing activity, each serum tested with SARS-CoV-2pp was tested in parallel with RD114pp (Figure S1), *i.e*., pseudoparticles coated with the glycoprotein gp70 of the endogenous feline retrovirus RD114 [15].

We first compared the neutralizing activity of the sera or plasma from our cohort of COVID-19 patients using both live virus and pseudoparticle assays. For live virus, the neutralization was assessed by ID50 (serum dilution that inhibits 50% of the infectivity) whereas for pseudoparticle assays, it was expressed as % of neutralization at the serum dilution of 1/100, both relative to a no serum condition. The nAb activity of each serum sample from the 140 patients as well as negative sera, *i.e*. sera that were collected before the emergence of the outbreak and sera from patients previously infected with other coronavirus (see below), were blindly quantified using these two detection methods. Similar results were obtained (Figure 1A), as indicated by the high Spearman’s rank correlation (rho=0.75). Hence, neutralization assays based on SARS-CoV-2 pseudoparticles can be reliably used to quantify nAb activity.

**Figure 1.**
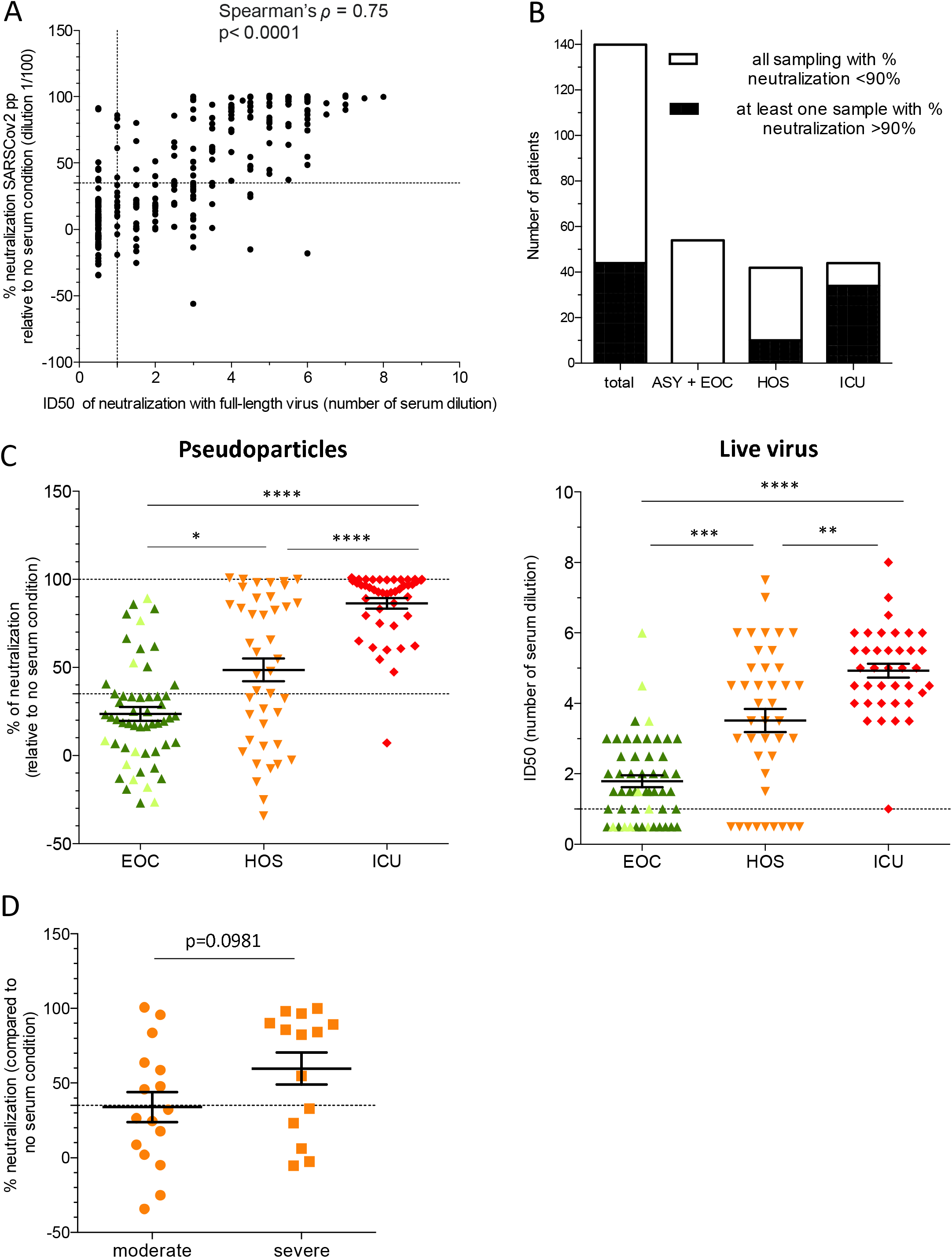
Serum neutralization of SARS-CoV-2 is correlated with the units of hospitalization of COVID-19 patients. (**A**) Correlation between ID50 of live virus, plotted as number of dilutions (two-fold dilutions, starting from serum diluted at 1/10), with percentage of neutralization of SARS-CoV-2pp for all tested samples. (**B**) Number of patients classified in the indicated groups according to the percentage of neutralization. In white: samples that induced a percentage of neutralization below 90%. (**C**) Comparison of the percentage of neutralization with SARS-CoV-2pp (left) or ID50 with live virus (right) for each patient classified according to the unit of hospitalization. For patients with serial serum samples, the sera collected at the closest to twenty days post-onset of symptoms were chosen. In light green are represented the asymptomatic patients (ASY) among the EOC patients. (**D**) Percentage of neutralization according to severity of symptoms in HOS patients.

### SARS-CoV-2 nAb response is correlated with COVID-19 symptoms severity

To explore potential humoral protective immune responses generated after SARS-CoV-2 infection, we compared the nAb activity of serum samples from patients in function of the severity of their symptoms. The nAb activity of their serum samples was evaluated using both SARS-CoV-2pp and wild-type virus. The cut-off for neutralization, *i.e*., 35%, was set using the mean neutralization of a 1/100 dilution of the negative sera + 2 standard deviation [19] for SARS-CoV-2pp. For wild-type virus, the cut-off for the ID50 was placed at the 1/10 dilution (first tested dilution) as all negative sera were found below this threshold.

We found that most patients from the different groups developed nAbs, although the neutralizing activity was highly variable (Figure 1C). Moreover, 44 patients (31.4%) exhibited a robust neutralization allowing over 90% neutralization of SARS-CoV-2pp (Figure 1B, total).

Strikingly, we found that ICU patients were particularly prone to display high nAb activity as compared to other groups with milder disease symptoms, such as HOS and EOC patients (Figure 1C). Indeed, only one patient in ICU did not develop a nAb response at the time of sampling, whereas for the HOS and EOC patients, the nAb activity was lower and more heterogenous. Specifically, 21.9% and 25% of patients in HOS and EOC categories respectively, did not develop nAbs at the time of sampling (Figure 1C - right). In addition, 34.5% of HOS patients had serum with no neutralizing activity at 1/100 and this number was increased to 70.7% in EOC patients (Figure 1C - left).

Concerning the HOS patients, we sought to classify the patients according to the severity of their symptoms. Accordingly, we defined as ‘severe’, the patients who had a respiratory rate >30/min and/or blood oxygen saturation <92% and/or lung lesions observed by CT scan and/or intensive oxygen therapy. The HOS patients were classified as ‘moderate’ if they did not fulfill the above criteria. However, we did not find a significant correlation (ρ=0.0981) between the severity of the symptoms of the HOS patients and their nAb titers, even though the % neutralization was higher in serum of patients with more severe disease forms (Figure 1D). This indicated that the severity of the symptoms may not be the only factor that explains the diversity of nAb activity in the HOS patients. Additionally, we found that neither the age and gender criteria nor the Ct of the first positive RT-qPCR titers (or the timing of sampling for qPCR) were associated with stronger or weaker neutralization (Table S1). This excluded an impact of the viral load or a bias due to the variability of the set-up of sampling in the heterogeneity of nAb activity observed in these patients.

Overall, these results indicated that the nAb activity was highly correlated with the severity of the symptoms, suggesting either that a robust humoral response was generated only in patients with severe symptoms or that the humoral response may contribute to the aggravation of the disease.

### SARS-CoV-2 anti-N and anti-S Abs correlate with neutralizing response

The high number of EOC patients without detectable nAb activity raises the question of the identification of correlates of protection in infected patients, since high titers of nAbs are generally thought to confer a protection against infection. Thus, we sought to identify whether IgG responses raised against components of SARS-CoV-2 particles could be correlated with the nAb activity detected in patients’ samples. For this, the anti-S and anti-N IgG responses were measured by two commercially-available tests. We found that most samples from SARS-CoV-2 positive patients were positive for at least one of either assay (Figure 2A, 2B). Interestingly, while anti-S IgG titers were highly correlated with the nAb titers (Spearman’s *ρ* = 0.7075, Fig 2B), the correlation was lower between anti-N IgG and nAbs (Spearman’s *ρ* = 0.5765, Figure 2A). Such differences were particularly obvious when we compared the distribution of the IgG among each category of patients. Indeed, while we found a low overlay in the profiles of anti-S IgG between EOC and ICU patients, which appeared similar to those of nAbs (compare Figure 1C vs. Figure 2C left), we detected a strong overlay in the value of anti-N IgGs between EOC and ICU patients (compare Figure 1C *vs*. Figure 2C right). This suggested that anti-S IgG values are more relevant to mark the presence and levels of nAb activity. Indeed, our results indicate that 95% of sera with above 124 AU/mL of anti-S IgGs were able to strongly neutralize SARS-CoV-2pp (>90% neutralization).

**Figure 2.**
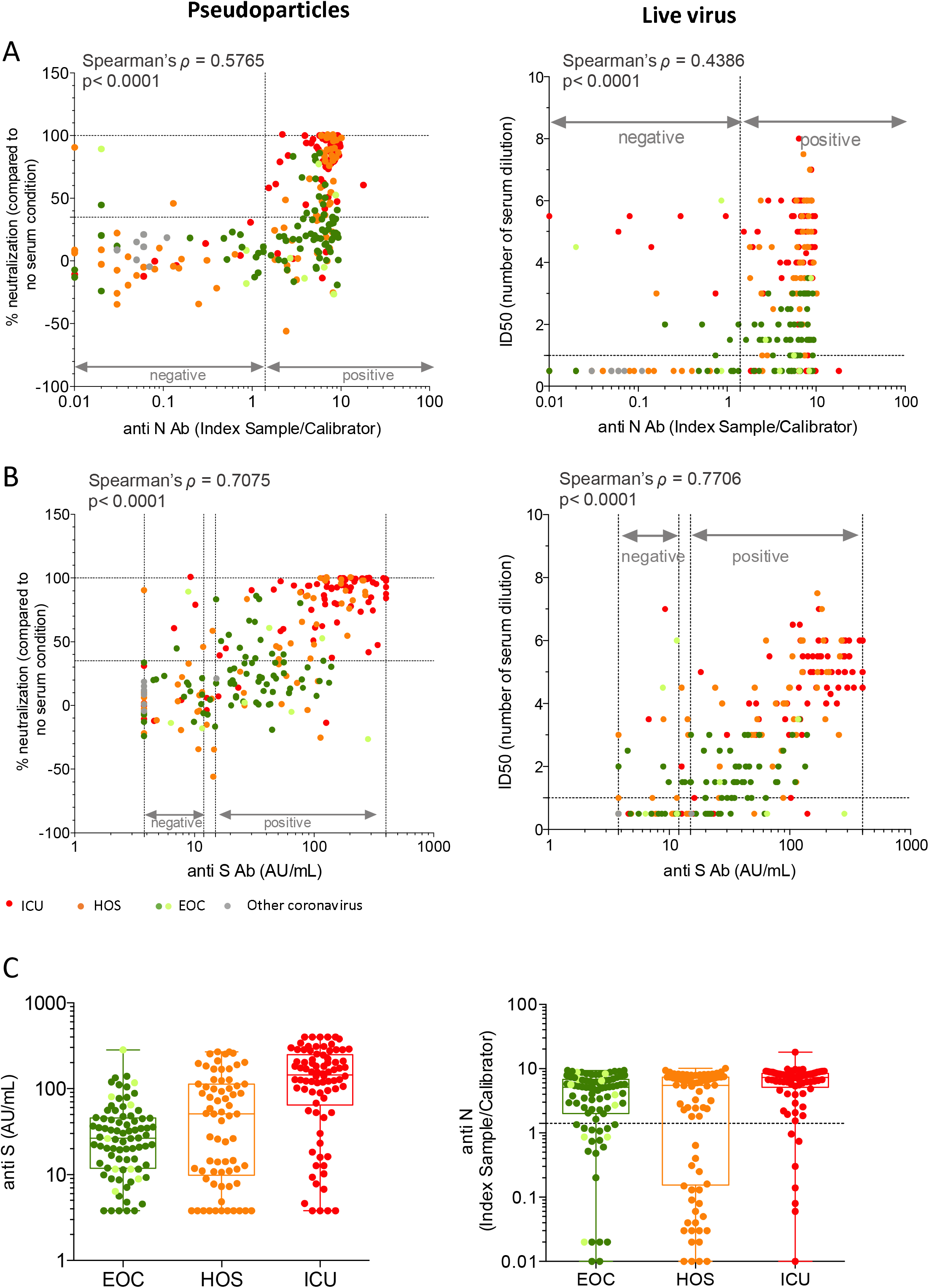
Serum neutralization of SARS-CoV-2 is correlated to anti-S antibodies. **(A)** Correlation between the % of neutralization of SARS-CoV-2pp (left) or ID50 of live virus (right) with seroconversion measured by anti-N antibodies. **(B)** Same as (A) with anti-S antibodies. **(C)** Anti-S (left) and anti-N (right) IgG values distribution in the three groups of patients.

Overall, these results indicated that the anti-S IgG response could be used as a surrogate of neutralizing activity in individuals.

### SARS-CoV-2 Ab kinetics in patient indicate a rapid waning of nAb titers

Next, we evaluated the kinetics of the anti-S IgG and nAb activities in our cohort of COVID-19 patients. Indeed, beside the diversity of clinical forms, one key feature of our cohort is the serial serum sampling for most patients, allowing to estimate the persistence of humoral factors. Both anti-S IgGs and nAb activities were generally detectable at 5-7 days post-onset of symptoms in patients who developed a humoral response, and rapidly increased to reach a peak before declining (Figure 3A, 3B). Hence, the kinetics of nAb titers as well as of anti-S IgG in each group were modelized with a second order polynomial (Figure 3A and 3B), which proved our best model among different ones tested (see parameters indicated in Figure 3). This regression analysis indicated that, instead of plateauing after the peak, the nAb activity could rapidly decrease, with an estimated half-life of 26 days. Additionally, the general tendency for anti-S IgGs seemed to follow the same pattern, with a peak and half-life similar to those calculated for the nAb activity, further confirming the correlation between anti-S IgG and neutralizing activity.

**Figure 3.**
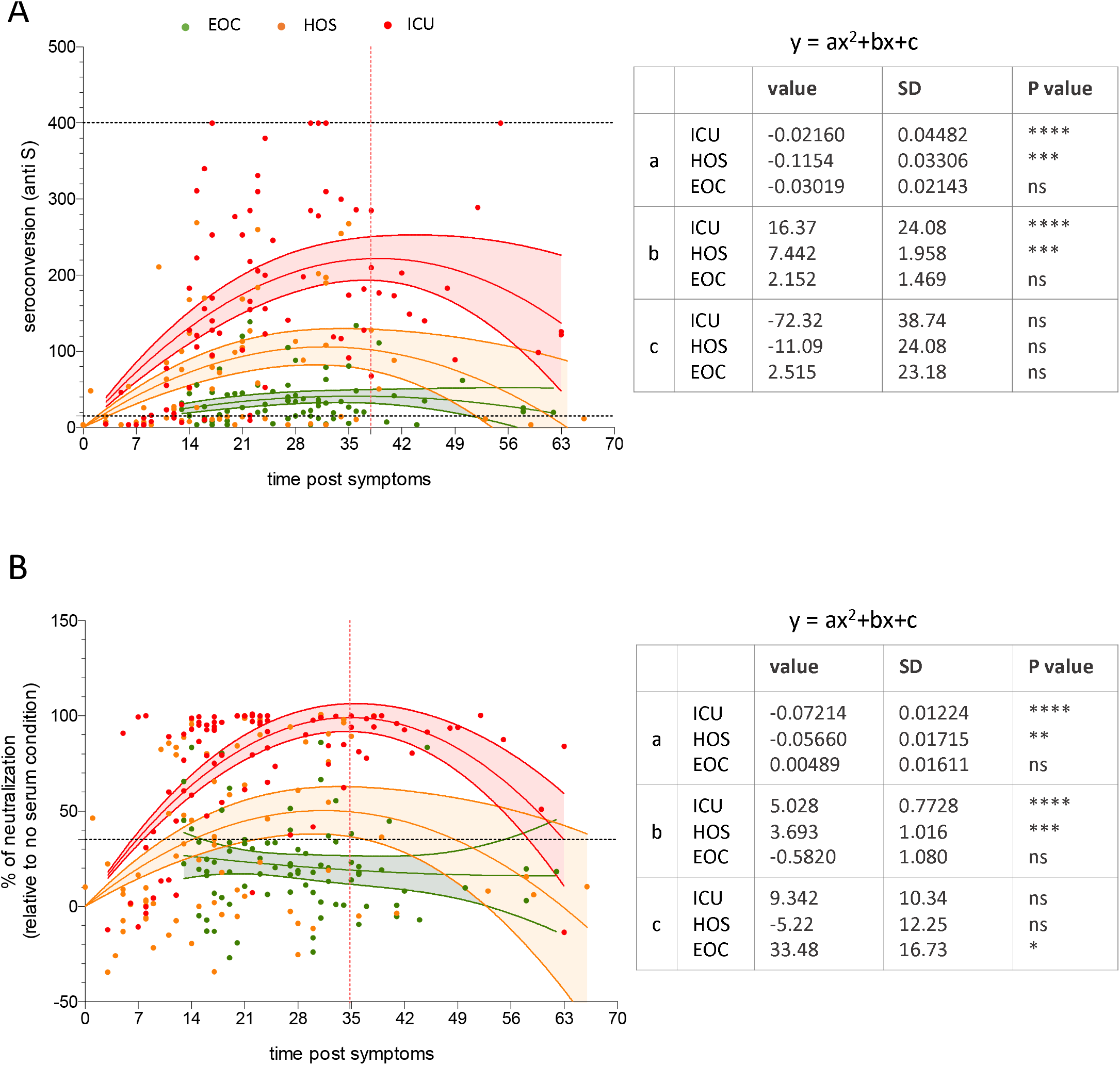
Apparition and longevity of neutralizing antibodies. **(A)** Seroconversion assessed by anti-S IgGs plotted against the days post-symptoms at which samples were collected. **(B)** Percentage of neutralization of each serum assessed with SARS-CoV-2pp plotted against the days post-symptoms at which samples were collected. The lines show the mean values expected from a second order polynomial regression, the ribbons indicate the pointwise 95% confidence intervals. The parameters of the regression analysis are indicated to the right.

### No cross-neutralization of SARS-CoV-2 can be induced by previous infection with alternative coronaviruses

Previous reports suggested that immunity against other coronaviruses may confer a certain degree of protection against alternative coronaviruses [20]. Hence, we explored the possibility that serum specimens from individuals diagnosed for OC43, 229E, NL63 and HKU1 coronaviruses (Figure 4A) but not infected with SARS-CoV-2 could cross-neutralize SARS-CoV-2. However, we found that none of the tested samples showed neutralizing activity above the cut-off of detection (Figure 4B), suggesting the absence of cross-neutralization between SARS-CoV-2 and endemic coronaviruses.

**Figure 4.**
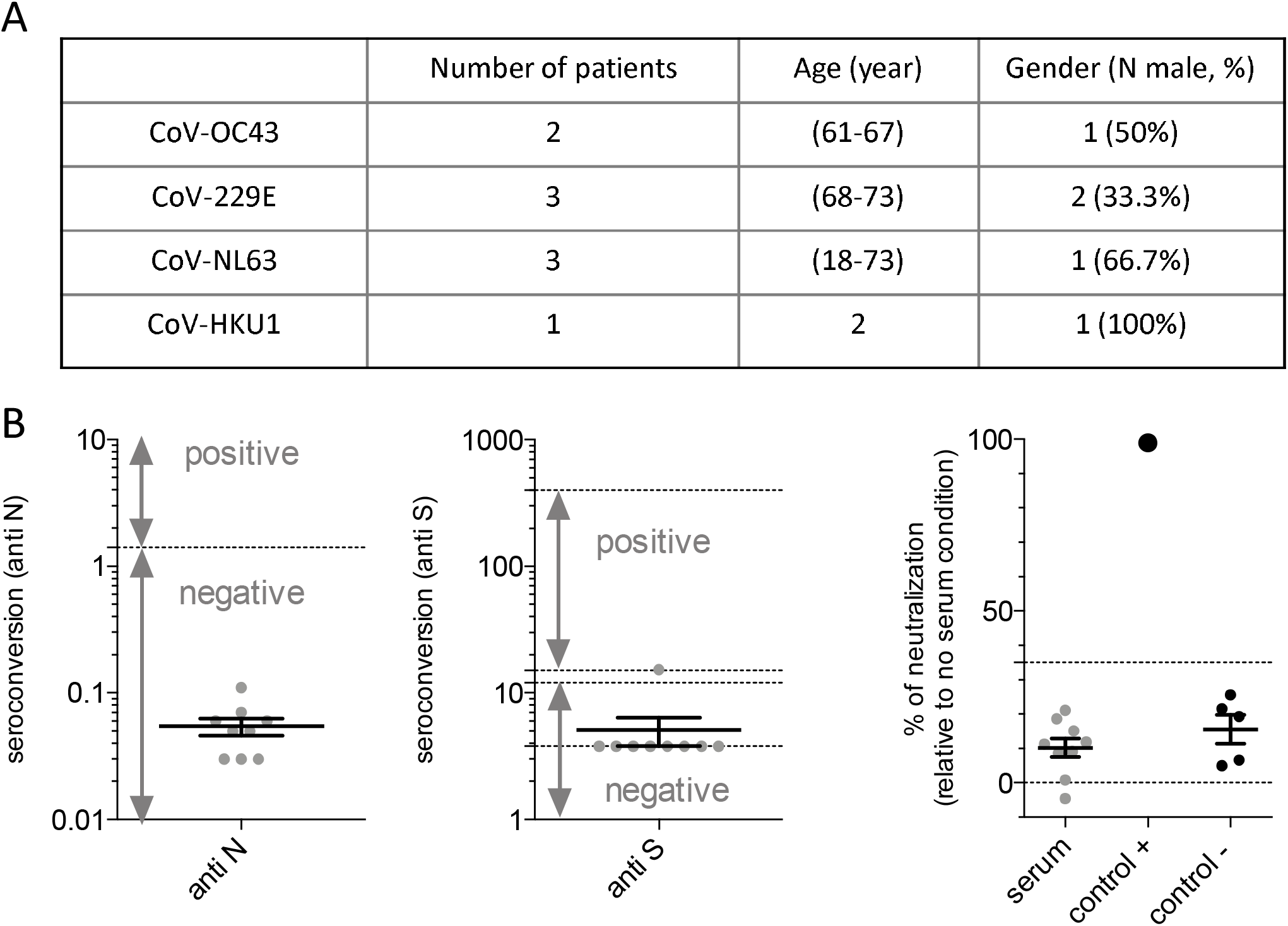
Sera from patients infected by endemic coronaviruses have no cross-neutralizing activity against SARS-CoV-2. **(A)** Characteristics of samples from patients infected with other coronaviruses. **(B)** Seroconversion assessed by anti-N (left), anti-S (middle) of SARS-CoV-2 or neutralization measured by SARS-CoV-2pp (right). For neutralization assays, a commercial anti-S antibody was used as positive control (control +) and five pre-pandemic serum samples were used as a negative control (control -).

### D614G substitution is not associated with resistance to SARS-CoV-2 neutralization

Considering the level of occurring mutations in coronaviruses, concerns have been raised about the emergence of SARS-CoV-2 immune escape mutants. In particular, the G614 variant in the Spike protein has progressively emerged and replaced the D614 initially present in the Wuhan strain to become the dominant pandemic form, which currently constitutes >97% of isolates world-wide [13]. To address the possibility of a neutralization escape phenotype potentially conferred by the D614G mutation, we used the SARS-CoV-2pp assay, which is particularly suitable to compare the nAb activity of serum specimens against pseudoparticles harboring or not this mutation. Yet, we found that that D614G mutation did not affect nAb activity of the serum samples from our cohort, as shown by similar profiles of neutralization (Figure 5), indicating that this highly prevalent mutation does not play a role in SARS-CoV-2 immune escape but rather may modulate viral fitness and infection [21].

**Figure 5:**
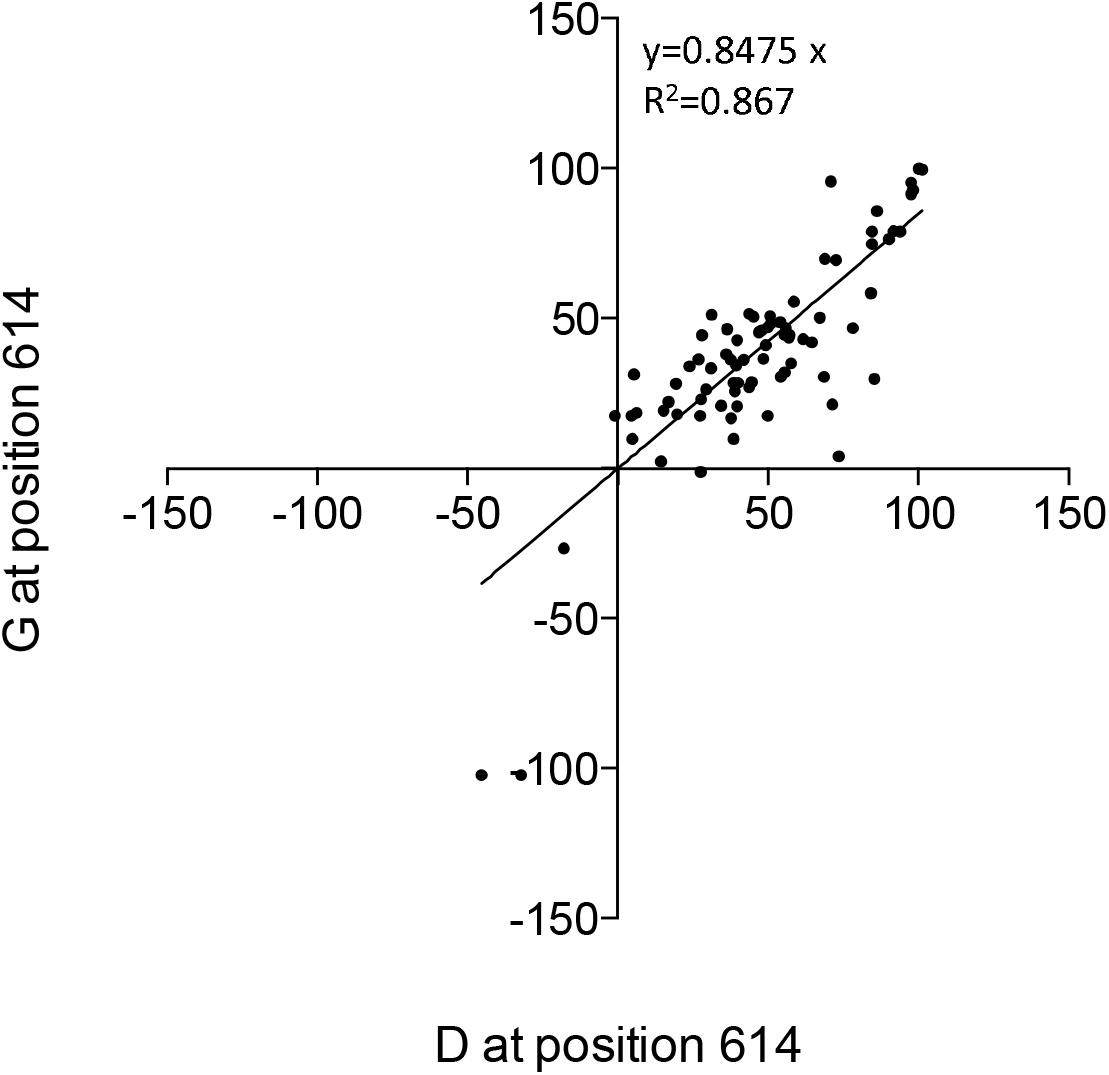
Residue at position 614 of SARS-CoV-2 Spike does not influence the activity of nAb. Percentage of neutralization of SARS-CoV-2pp using Spike protein with either a G or D at position 614.

## Discussion

Our understanding of the nature of protective immune response to SARS-CoV-2 is currently limited but is likely to involve both cellular and humoral immunity. Here, we characterize some features of the humoral response in a cohort of individuals infected with SARS-CoV-2. The serum specimens used in this study were collected according to the routinely clinical management, without selection of specific criteria. Although this may limit its statistical interpretation, this cohort represents a particularly appropriate picture of the different forms of severity for COVID-19 patients and, thus, allows an original longitudinal study.

Using either a live virus or a pseudotyped system, we compared the nAb response in sera from patients with different levels of COVID-19 severity, according to their hospitalization status, including patients in intensive care unit but also SARS-CoV-2-positive individuals with more moderate disease forms. Recent reports have evaluated the neutralizing activity of sera from COVID-19 patients with controversial results. While a study suggested that patients rapidly develop neutralizing antibodies [22], other studies indicated that the humoral response [23] and nAb activity [24, 23] are correlated to several parameters including the severity of the disease, resulting in the absence of detection of nAb activity in a significant number of mild symptomatic and asymptomatic patients [25]. However, such assumptions were based on a very limited number of observations, since those studies included less than 10 patients with the most severe forms. Our cohort is composed of 140 patients, one third of whom are ICU patients, which represent the largest clinical cohort published to date in which neutralizing activity has been evaluated. Those characteristics allowed us to address important questions such as correlations of nAb activity with disease severity, IgG response, or kinetics of antibody levels. Here, we confirmed the heterogeneity of the humoral response in COVID-19 patients. While we found that parameters, such as age or gender, were not associated with nAb activity, we show that among patients with mildest disease forms, many did not exhibit robust neutralizing activities (Figure 1). Importantly, we demonstrated that the severity of the disease, as assessed by the hospitalization unit status, strongly correlated with the levels of nAbs as well as with anti-S IgG titers (Figure 1 and Figure 2). This ties with a recent observation that the strongest T cell signals are found in patients with the most severe disease forms, raising the question of the beneficial vs. detrimental effect of T cell activation in SARS-CoV-2 pathogenesis [26].

Importantly, while for most viral infections, high neutralizing antibody titers are usually associated with viral clearance, it seems that a robust nAb activity does not confer protection against disease progression in COVID-19 patients. One possibility is that disease severity is correlated with higher viral loads and hence, with more antigens available for induction of antibodies. Yet, no correlation between viral loads and nAb activity could be identified in our cohort (Table S1), though the assessment of the viral loads by the first positive qPCR could be unreliable, due to parameters that can influence the quality of sampling [27]. Alternatively, a robust humoral response may be a feature of an overall exaggerated immune activation in severe SARS-CoV-2 infection. Indeed, antibodies could mediate additional immune functions that may have both protective and pathological consequences. In addition, humoral responses have been shown to be corelated with cytokines and chemokines levels in COVID-19 patients [28] that are the main effectors of severe systemic inflammatory responses known as “cytokine storm” found in the most severe forms of the disease [29]. Thus, an uncontrolled humoral response may also contribute directly to the pathogenesis of the disease by promoting organ damages, but this mechanism has to be demonstrated for SARS-CoV-2. In this perspective, questions about the use of convalescent plasma transfusion as a treatment for COVID-19 could be raised, since it is uncertain whether such transfusion may lead to an aggravation of the disease [30].

The correlation between seroconversion and nAb activity was also analyzed (Figure 2 and Figure 3). As expected, both anti-S and anti-N IgG levels correlated with nAb titers, although correlation with anti-S antibodies was the strongest, which likely reflects Spike being the main target of nAbs for diverse coronaviruses, including SARS-CoV-2 [31]. Thus, should neutralizing activity be associated with protection against subsequent re-infections, the determination of anti-S IgG titers, by *e.g*., ELISA, may be useful to discriminate protected from unprotected individuals, in particular for patients with the most moderate forms, since they are less prone to develop robust neutralizing activity. In our cohort, 95% of sera with anti-S antibody values above 124 AU/mL were associated with robust neutralization (90% neutralization or more), suggesting that anti-S antibody determination could be used as an evaluation of nAb activity, rather than for anti-N IgG assays.

We also addressed the question of the stability of the nAb levels, owing to the availability of several serial samples for most of the patients used in the study. We confirmed a tendency for a decrease of the nAb activity as well as of anti-S IgG after reaching a peak, as previously shown by others [32], indicating that for some patients, the nAb activity may be very transient (Figure 3). Importantly, for ICU patients, we were able to give a more precise estimation of the persistence of the neutralizing activity, with a preliminary estimation of 26 days for its half-life. Thus, our model predicts that the SARS-CoV-2 nAb activity is likely to rapidly vanish. These results, which need to be confirmed by studying alternative COVID-19 cohorts and more patients, are in sharp contrast with SARS-CoV-induced disease, for which the nAbs could be detected for about 270 days with ID50>100 [33]. Thus, for SARS-CoV-2, it remains to be determined if such waning of the nAb activity is associated with an absence of protection or, alternatively, if potential re-infections may trigger immune memory and induce faster and higher production of nAbs, eventually leading to a better control of the infection and/or to reduced symptoms. Further studies are warranted for the evaluation of the persistence of SARS-CoV-2 nAbs.

Naturally occurring variants in S proteins have been reported since the beginning of the pandemic. Among them, a variant with a single mutation in 614 position (D614G) has become the currently dominant circulating virus. This variant has been associated with increased infectivity though it did not exhibit resistance to nAb present in convalescent sera [21]. Our report confirms that, by comparing the neutralizing activity for SARS-CoV-2pp with a D614 and G614 S, this mutation is not associated with resistance to neutralization.

Finally, we also analyzed 9 serum samples from patients infected with alternative coronaviruses that cause mild symptom in adults, including respiratory illnesses, and enteric and neurological diseases [34]. The cross reactivity between other coronaviruses and SARS-CoV-2 immunity is key to understand, as it might influence the severity of the disease or the response to a vaccine [35]. In line with this, T cell reactivity against SARS-CoV-2 was observed in unexposed people [36]. Interestingly, by investigating the cross reactivity of circulating Abs, we found that none of the 9 samples displayed cross-reacting nAbs against SARS-CoV-2 infection. Furthermore, cross-neutralization against SARS-CoV-2 can be induced by sera from convalescent SARS-CoV patients, which is likely due to high homology between these two viruses, although it seems to be serum-dependent [37, 38]. Thus, our results indicate an absence of cross-neutralization between SARS-CoV-2 and other endemic human coronaviruses, suggesting that previous infections with alternative coronaviruses do not protect against SARS-CoV-2 infection by generation of nAbs.

## Data Availability

All data are available upon request

## Acknowledgments

We thank D. Lavillette for providing the SARS-CoV-2 Spike expression vector and B. La Scola for providing a clone of VERO-E6 cells.

We acknowledge the contribution of SFR Biosciences (UMS3444/CNRS, US8/Inserm, ENS de Lyon, UCBL) ANIRA-Cytometry facility for excellent technical assistance and support.

The laboratory of FLC received financial support from the LabEx Ecofect (ANR-11-LABX-0048) of the ‘‘Université de Lyon”, within the program ‘‘Investissements d’Avenir” (ANR-11-IDEX-0007) operated by the French National Research Agency (ANR), the ANR (grant from RA-Covid-19), the Fondation pour la Recherche Médicale (FRM), and Inserm Transfert.

## Authorship Contributions

Study concept and design: BB, MV, TB, FLC, SD, VL, BP. Inclusion and characterization of patients: PB, EBN, TG, CP, MV, TB. Acquisition of data: BB, SD, VL, SP, SG, JR, PV, BP.

Analysis and interpretation of data: OA, FG, BB, SP, TB, FLC, SD, VL, BP. Drafting of the manuscript: BB TB, FLC, SD, VL, BP. Funding acquisition: BB, TB, FLC, SD, VL, TW.

## Disclosure of Conflicts of Interest

The authors have declared that no competing interests exist.

## Supplementary Material

**Figure S1 legend. Neutralizing antibodies are specific to SARS-CoV-2** Results of neutralization using control of pseudoparticles generated with the RD114 cat endogenous virus surface glycoprotein.

**Table S1.** Absence of correlation between age or the gender criteria or Ct of first PCR or timing of sampling with neutralization.

| Criteria | P-value |
| --- | --- |
| **ICU** |  |
| Age | 0.694 |
| Gender | 0.367 |
| Ct 1<sup>st</sup> PCR | 0.843 |
| Timing of sampling | 0.216 |
| **HOS** |  |
| Age | 0.086 |
| Gender | 0.44 |
| Ct 1<sup>st</sup> PCR | 0.8 |
| Timing of sampling | 0.97 |
| **EOC** |  |
| Age | 0.92 |
| Gender | 0.51 |
| Ct 1<sup>st</sup> PCR | 0.14 |
| Timing of sampling | 0.65 |

## Notes

### Competing Interest Statement

The authors have declared no competing interest.

